# Hebbian plasticity induced by temporally coincident BCI enhances post-stroke motor recovery

**DOI:** 10.1101/2023.09.28.23296226

**Authors:** Johanna Krueger, Richard Krauth, Christoph Reichert, Serafeim Perdikis, Susanne Vogt, Tessa Huchtemann, Stefan Dürschmid, Almut Sickert, Juliane Lamprecht, Almir Huremovic, Michael Görtler, Slawomir J. Nasuto, I-Chin Tsai, Robert T. Knight, Hermann Hinrichs, Hans-Jochen Heinze, Sabine Lindquist, Michael Sailer, Jose del R. Millán, Catherine M. Sweeney-Reed

**Author notes:** Corresponding author: Catherine Sweeney-Reed.

## Abstract

Functional electrical stimulation (FES) can support functional restoration of a paretic limb post-stroke. Hebbian plasticity depends on temporally coinciding pre- and post-synaptic activity. A tight temporal relationship between motor cortical (MC) activity associated with attempted movement and FES-generated visuo-proprioceptive feedback is hypothesized to enhance motor recovery. Using a brain–computer interface (BCI) to classify MC spectral power in electroencephalographic (EEG) signals to trigger FES-delivery with detection of movement attempts improved motor outcomes in chronic stroke patients. We hypothesized that heightened neural plasticity earlier post-stroke would further enhance corticomuscular functional connectivity and motor recovery. We compared subcortical non-dominant hemisphere stroke patients in BCI-FES and Random-FES (FES temporally independent of MC movement attempt detection) groups. The primary outcome measure was the Fugl-Meyer Assessment, Upper Extremity (FMA-UE). We recorded high-density EEG and transcranial magnetic stimulation-induced motor evoked potentials before and after treatment. The BCI group showed greater: FMA-UE improvement; motor evoked potential amplitude; beta oscillatory power and long-range temporal correlation reduction over contralateral MC; and corticomuscular coherence with contralateral MC. These changes are consistent with enhanced post-stroke motor improvement when movement is synchronized with MC activity reflecting attempted movement.

## Introduction

Stroke is a leading cause of motor disability^1^, with upper limb impairment occurring in over 75% of patients following acute stroke^2^. Despite reductions in mortality and morbidity through thrombolytic therapy, a third or less of patients meet the criteria, and over half of those receiving it are left with functional deficits^3^. Motor recovery depends on neural plasticity and the reorganization of structural and functional motor networks to re-establish corticomuscular connectivity^4–8^. Neural plasticity is task-specific, time-dependent, and environmentally-influenced^9^. Various approaches to re-establishment and reinforcement of connectivity between paretic musculature and residual motor areas are based on targeting Hebbian plasticity by synchronizing movement-associated visuo-proprioceptive feedback and motor cortical electrophysiological correlates of movement within a narrow time window^10,11^. Functional electrical stimulation (FES) is an established therapeutic tool for assisting movement attempts and promoting motor recovery. Studies involving chronic and subacute stroke patients have shown enhanced motor recovery when FES delivery is temporally coupled to movement attempts detected in brain electrical activity, using a brain–computer interface (BCI)^12–14^. Electroencephalographic (EEG) signals recorded over motor cortex provided the input to a classifier, and FES was triggered when features derived from these signals were classified as reflecting attempted movement as opposed to rest. Although starting rehabilitation early post-stroke is associated with better motor outcomes, putatively due to heightened neural plasticity^4,15,16^, the majority of studies implementing BCI-FES-based rehabilitation focus on patients in the chronic phase^17,18^. We hypothesized that earlier initiation of BCI-FES would improve corticomuscular functional connectivity, resulting in greater motor recovery. Functional connectivity here refers to restoring dependency of muscle contraction on motor cortical activity. Dependency is reflected in movement occurring on voluntary motor cortical activity modulation, which we aimed to support through BCI-FES, and in increased statistical dependency between EEG and movement-related electromyographic (EMG) activity, which can be indexed by corticomuscular coherence (CMC)^7,19,20^. We also performed an exploratory evaluation of neural correlates of motor recovery in patients receiving BCI-FES to gain a better understanding of potential mechanisms of action. The early phase post-stroke poses challenges in therapy program completion, and heterogenous patient groups with cortical and subcortical stroke, affecting either hemisphere, are commonly included. Here we compared outcomes in a BCI-timed (BCI-FES) and a randomly timed (Random-FES) group in a matched lesion subgroup from the Magdeburg patient cohort (German Clinical Trials Register: DRKS00007832; DRKS00011522). BCI-FES and Random-FES patients had suffered a subcortical stroke affecting the non-dominant hemisphere, and the tight uniformity of the study group enabled group-level comparisons of electrophysiological and behavioral markers over the treatment period.

While clinical outcome is the primary focus in evaluating rehabilitation measures, understanding the mechanisms underlying recovery is the key to informing further development. Electrophysiological and functional measures of brain activity can provide potential markers of modulation during therapy. Brain oscillatory activity^21^ and corticomuscular functional connectivity^20^ have been proposed as biomarkers of post-stroke recovery. Here we compared clinical outcome and neural correlates of motor recovery in patients in the acute and subacute phases post-stroke allocated to BCI-FES or FES delivered without a tight temporal relationship with EEG correlates of movement attempts (BCI- and Random-FES groups). The patients underwent a three-week FES rehabilitation program, with transcranial magnetic stimulation (TMS)-induced motor evoked potential (MEP) amplitude measurement as a part of their routine clinical evaluation, and high-density EEG recordings for the purpose of the study. The EEG analyses included sensorimotor cortical spectral power, CMC, and long-range temporal correlation (LRTC).

In the BCI-FES group, movement attempts were detected by online classification of EEG signals. The sensorimotor rhythm refers to oscillations in brain electrical activity over motor cortical regions in the alpha (8-12 Hz) and beta (13-30 Hz) frequency ranges. Event-related desynchronization and synchronization (ERD/ERS) index reduction/increase of the sensorimotor rhythms, detectable as changes in EEG starting before and changing over the course of movement^22,23^. They provide well-established indices of actual movement, as well as of imagined movement^24^ and movement attempts^25^ and are commonly used in BCIs^26^. EEG was recorded from each patient during a training session of cued movement attempts and rest periods. The electrode locations and frequencies at which oscillatory power differences were greatest between movement and rest were selected as features for classifier training.

We focused our analyses on the largest possible uniform patient group, due to the importance of laterality in post-stroke recovery: right-handed patients with a non-dominant hemisphere stroke. Handedness has an impact on movement- and imagined-movement-related sensorimotor cortical oscillatory activity and fMRI activation in healthy participants^27–29^, and activation patterns during post-stroke rehabilitation differ according to whether the dominant or non-dominant hemisphere is affected^30,31^.—We examined electrophysiological changes over the treatment period both in contralesional and ipsilesional motor cortical regions. Shifts of abnormal bilateral motor area activation during paretic hand movement, in the subacute phase, toward a more unilateral activation pattern of ipsilesional motor areas in chronic stroke, is associated with better motor outcome^32,33^. While contralesional motor cortical activity is associated with poorer motor outcomes in the chronic phase post-stroke, this activity appears to play an important role early post-stroke^32^.

The primary outcome measure was change in the Fugl-Meyer Assessment of the Upper Extremity (FMA-UE) score from before to after treatment. We also examined potential neural correlates of a direct effect of BCI-FES on relevant neural processes. The amplitude of TMS-induced MEPs provides an index of the integrity of corticospinal pathways, and these were measured before and after the treatment program. Based on delivery of FES in temporal association with movement-associated spectral power changes in the sensorimotor rhythm, we compared spectral power across the alpha and beta frequency ranges after, with that before the treatment program, in each group. We compared at a group and an individual level and evaluated correlation between sensorimotor oscillatory power and FMA-UE score after treatment. As the aim was re-establishment of corticomuscular functional connectivity, we also assessed change in the EEG–EMG coherence in the same frequency range before with after treatment in each group^20^. We also evaluated a potential impact of BCI-FES on LRTC. LRTC provides an index of correlation between different time periods in a time series, reflecting the extent to which neuronal systems are at a near-critical state permitting rapid changes in functional connectivity as processing demands change over time^34^. LRTC is postulated to facilitate information transfer in neuronal networks, with physiological memory of a past activity influencing future activity through continuous modification and recurrent interactions between ongoing activity and stimulus-induced changes in activity^34,35^. Cumulative modification in network functional connectivity, due to activity-dependent plasticity, has been proposed to provide the physiological mechanism underlying the power law correlations in ongoing oscillatory neuronal network activity, influencing future recruitment of neurons to engage in particular oscillatory activity^34^. LRTC observed in EEG shows power-law behavior, suggesting similar underlying neurodynamic processes on different time scales^35^. The amplitude envelope of alpha and beta oscillations displays intermittent fluctuations and power-law decay of the autocorrelation over hundreds of seconds, suggesting a self-organized dynamical critical state^34^. Task-relevant neural assemblies, defined by temporal relationships between activity in different brain areas, form and dissolve over time^36,37^. Sensory stimuli result in reorganization of ongoing endogenous brain dynamics^38^. As activity is propagated through cortical networks, altering functional connectivity, reflected in changes in LRTC, and influencing future neuronal recruitment, somatosensory stimuli disrupt these transient neural assemblies, degrading ongoing LRTC^34^. We hypothesized that tight temporal coupling between motor cortical oscillatory power and the somatosensory stimulus in the BCI-FES group would result in a greater LRTC reduction than a somatosensory stimulus delivered independently of motor cortical activity corresponding to a movement attempt.

## Results

### Patients

Of the patients recruited in Magdeburg (N = 32), 62.5 % (n = 20) completed the rehabilitation program (Figure 1). The reasons for discontinuing participation were complete recovery (n = 2), finding the therapy too tiring (n = 1), the sequelae of a previously diagnosed psychiatric (n = 4) or physical illness (n = 4), and the patient leaving the region (n = 1). Ten patients were allocated to the BCI-FES group and 10 patients to the Random-FES group. The analysis was applied to the largest sub-group of patients with similar lesion location, which was those whose non-dominant hemisphere was affected by a subcortical stroke, resulting in equal BCI-FES (n = 6) and Random-FES (n = 6) group sizes.

**Figure 1.**
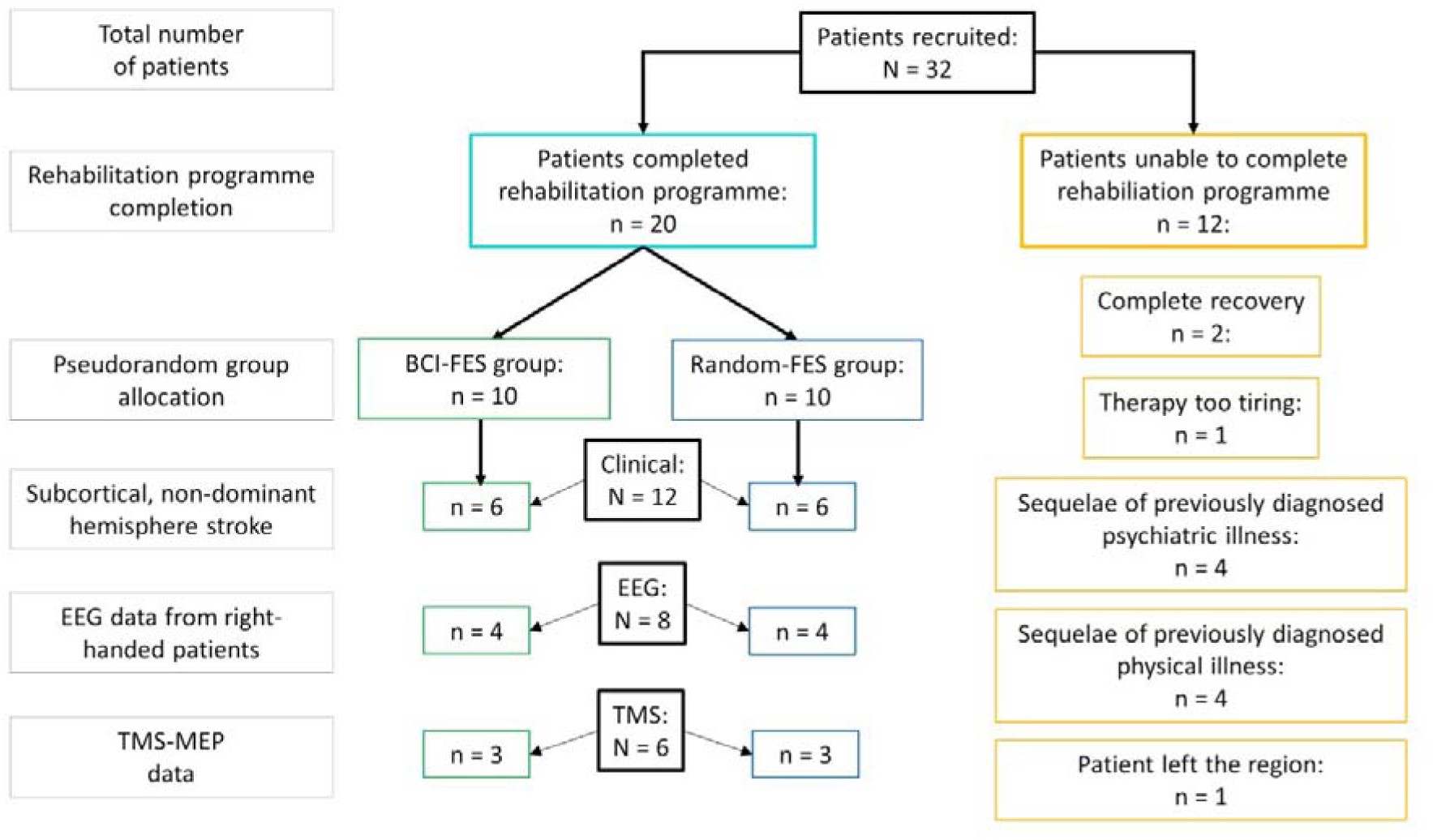
Numbers of patients in the groups receiving functional electrical stimulation timed according to a brain–computer interface (BCI-FES) and timed randomly (Random-FES) completing the rehabilitation program and included in each evaluation.

### BCI features

The features (electrode locations and spectral power frequencies) that were selected at each re-training of the classifier for the BCI-FES group patients changed over the course of treatment in all patients (Supplementary Fig. 1). Early in the program, bilateral features provided the best classification, with a tendency towards ipsilesional (contralateral) features being selected by the final training of the classifier. While the features included power in both the alpha and beta frequencies throughout, alpha power continued to be relevant by the end of the treatment period. By week 4 or later, all classifiers included an alpha power feature. Only one patient had an ipsilateral beta feature by the end.

### Clinical evaluation

Examining the FMA-UE scores before and after the program, an interaction was observed between *Time* and *Group* (F(1) = 8.03, p = 0.030; η_p_^2^ = 0.57) (Fig. 2). No other interactions were significant. A main effect of *Time* was also observed (F(1,6) = 8.93, p = 0.024; η_p_^2^ = 0.60). No other within-subject main effects were significant. No between-subject effects were significant. Post hoc pairwise comparisons showed a significant increase in FMA-UE score from pre- to post-treatment in the BCI-FES group (Pre: mean [M] = 11.3, standard deviation [SD] = 4.6; Post: M = 27.5, SD = 17.5; p = 0.004) but not in the Random-FES group (Pre: M = 10.0, SD = 3.6; Post: M = 14.8, SD = 12.4; p = 0.77). The scores did not differ between the groups pre-treatment (p = 0.81), and a trend towards a higher score in the BCI-than the Random-FES group was seen post-treatment (p = 0.062).

**Figure 2.**
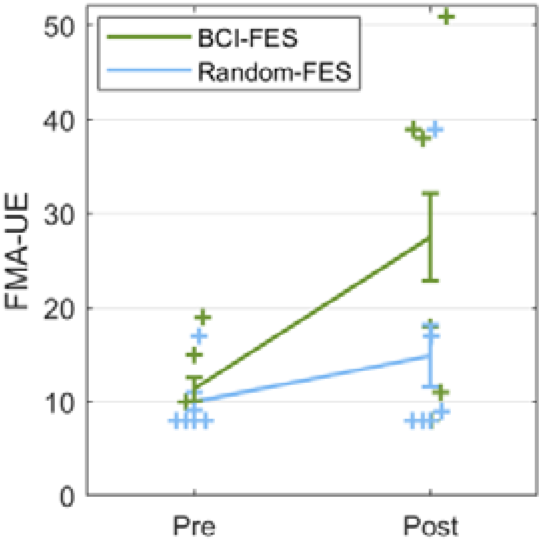
The Fugl-Meyer Assessment of the upper extremity (FMA-UE) showed greater motor recovery in the BCI-FES group post-treatment than the Random-FES group. Interaction between *Time* and *Group:* F(1) = 8.03, p = 0.030; η_p_^2^ = 0.57, correcting for covariates *Age*, *Sex*, *Days Post-Stroke*, and *Days of Therapy.* Post hoc tests pre- to post-treatment: BCI-FES: p = 0.004; Random-FES: p = 0.77. (The maximum score of the FMA-UE is 66 points.) Error bars = standard error of the mean.

Of the secondary clinical outcome measures, a *Group* x *Time* interaction (F(1) = 6.00, p = 0.043; η_p_^2^ = 0.52) and a main effect of *Time* (F(1) = 6.00, p = 0.041; η_p_^2^ = 0.53) were only observed for the National Institute of Health Stroke Scale (NIHSS) upper limb score. Post hoc testing showed a significant improvement in the BCI-FES (Pre: M = 3.2, SD = 1.2; Post: M = 1.7, SD = 1.6; p = 0.009) but not in the Random-FES group (Pre: M = 3.2, SD = 1.2; Post: M = 2.8, SD = 1.0; p = 0.92).

When *Therapy start* (Acute, Subacute) was included as a between-subject factor, the only significant interaction remained *Time* x *Group* (F(1) = 6.66, p = 0.049; η_p_^2^ = 0.57) (Fig. 3). Post hoc tests showed an increase in FMA-UE score in the BCI-FES group (p = 0.010) but not in the Random-FES group (p = 0.89). The FMA-UE score increased in the BCI-FES group from pre- to post-treatment when therapy was started in the acute (within one month of stroke: Pre: M = 12.3, SD = 5.9; Post: M = 33.3, SD = 20.4; p = 0.016) but not the subacute (one to six months post-stroke: Pre: M = 10.3, SD = 4.0; Post: M = 21.7, SD = 15.8; p = 0.21) phase. The increase was not significant in the Random-FES group, starting in either the acute (Pre: M = 9.0, SD = 1.7; Post: M = 11.3, SD = 4.9; p = 0.94) or the subacute phase (Pre: M = 11.0, SD = 5.2; Post: M = 18.3, SD = 17.9; p = 0.78).

**Figure 3.**
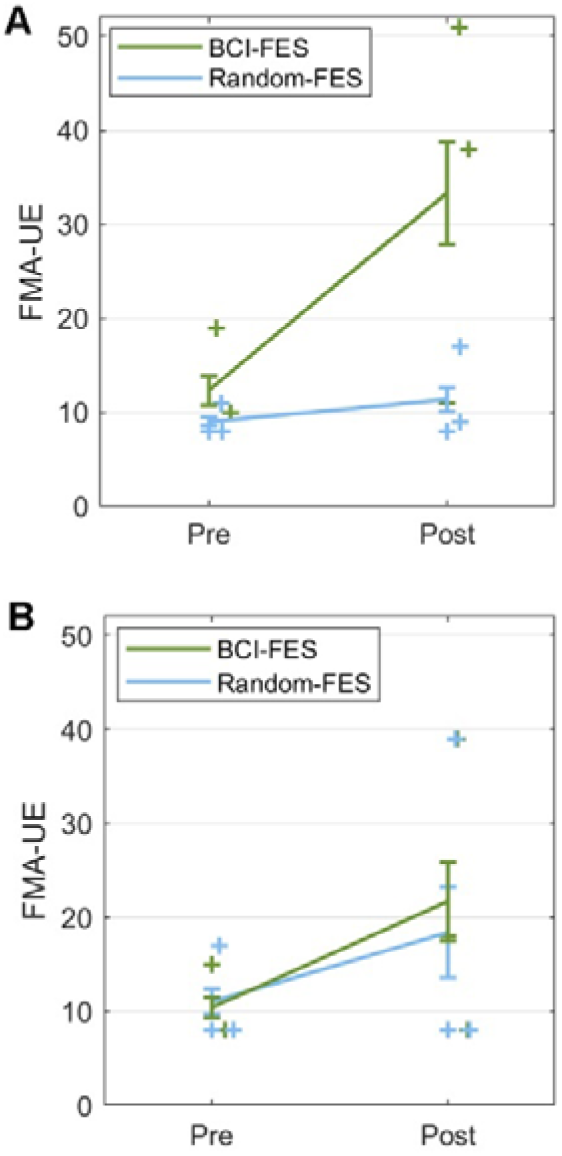
The improvement in Fugl-Meyer Assessment of the upper extremity (FMA-UE) score was greatest in the BCI-FES group from pre- to post-treatment in patients who started treatment in the acute phase (within one month) post-stroke compared with patients in the Random-FES group and with patients in either group starting treatment in the subacute phase. Including *Therapy start* as a factor: interaction *Time* x *Group* (F(1) = 6.66, p = 0.049; η_p_^2^ = 0.57) **A**. Patients starting treatment in the acute phase: post hoc p = 0.016. **B**. Patients starting treatment in the subacute phase: post hoc p = 0.78. Error bars = standard error of the mean.

### TMS

TMS measurements were available from patients with a subcortical stroke from both groups (BCI-FES: n = 3; Random-FES: n = 3). An interaction was observed between *Group* and *Time* (F(1) = 27.69, p = 0.034; η_p_^2^ = 0.93) (Fig. 4). There was no main effect of *Group* (F(1,2) = 9.12, p = 0.094) or *Time* (F(1,2) = 1.36, p = 0.36). Post hoc tests revealed a significant amplitude increase from pre- to post-treatment in the BCI-FES group (p = 0.012) only (Random-FES group: p = 0.50).

**Figure 4.**
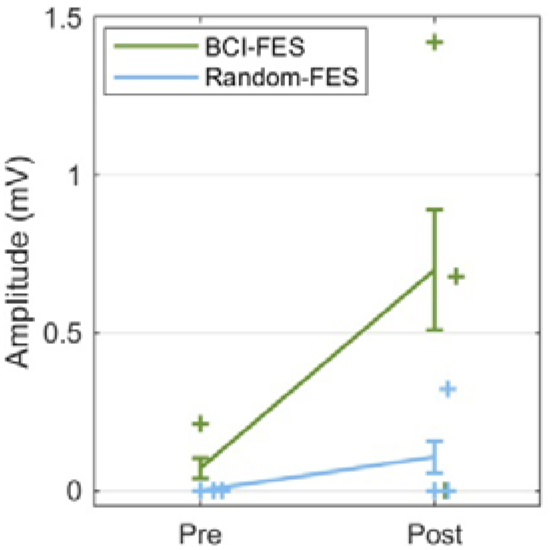
The amplitude of the transcranial magnetic stimulation (TMS)-induced motor evoked potentials increased following treatment in the BCI-FES group but not the Random-FES group. Interaction: *Group* and *Time* (F(1) = 27.69, p = 0.034; η_p_^2^ = 0.93), correcting for covariates. BCI-FES: post hoc p = 0.012; Random-FES: post hoc p = 0.050. TMS was applied at electrode location C2, contralateral to the affected limb. Error bars = standard error of the mean.

### High-density EEG

Oscillatory spectral power differed between pre- and post-treatment in the BCI-FES group (p = 0.036), with a reduction in lower beta (15-23 Hz) oscillatory spectral power around 0.5 to 1.5 s following the movement cue over the ipsilesional motor cortex (at electrode C2), which was not seen in the Random-FES group (Fig. 5). Spectral power was compared before and after treatment for each patient on the contralateral (C2) and ipsilateral (C1) side to movement, at the time post-movement at which the pre- to post-movement change was greatest (1.2 to 1.4 s) (Fig. 5). Beta power reduction over the treatment period was most consistent across individuals in the BCI-FES group over the ipsilesional motor cortex, contralateral to movement (Fig. 6). The contralateral beta power after therapy correlated with the FMA-UE score (r(2) = 0.96, p = 0.044) (Fig. 6). No significant correlation was observed in the Random-FES group nor in either group before therapy.

**Figure 5.**
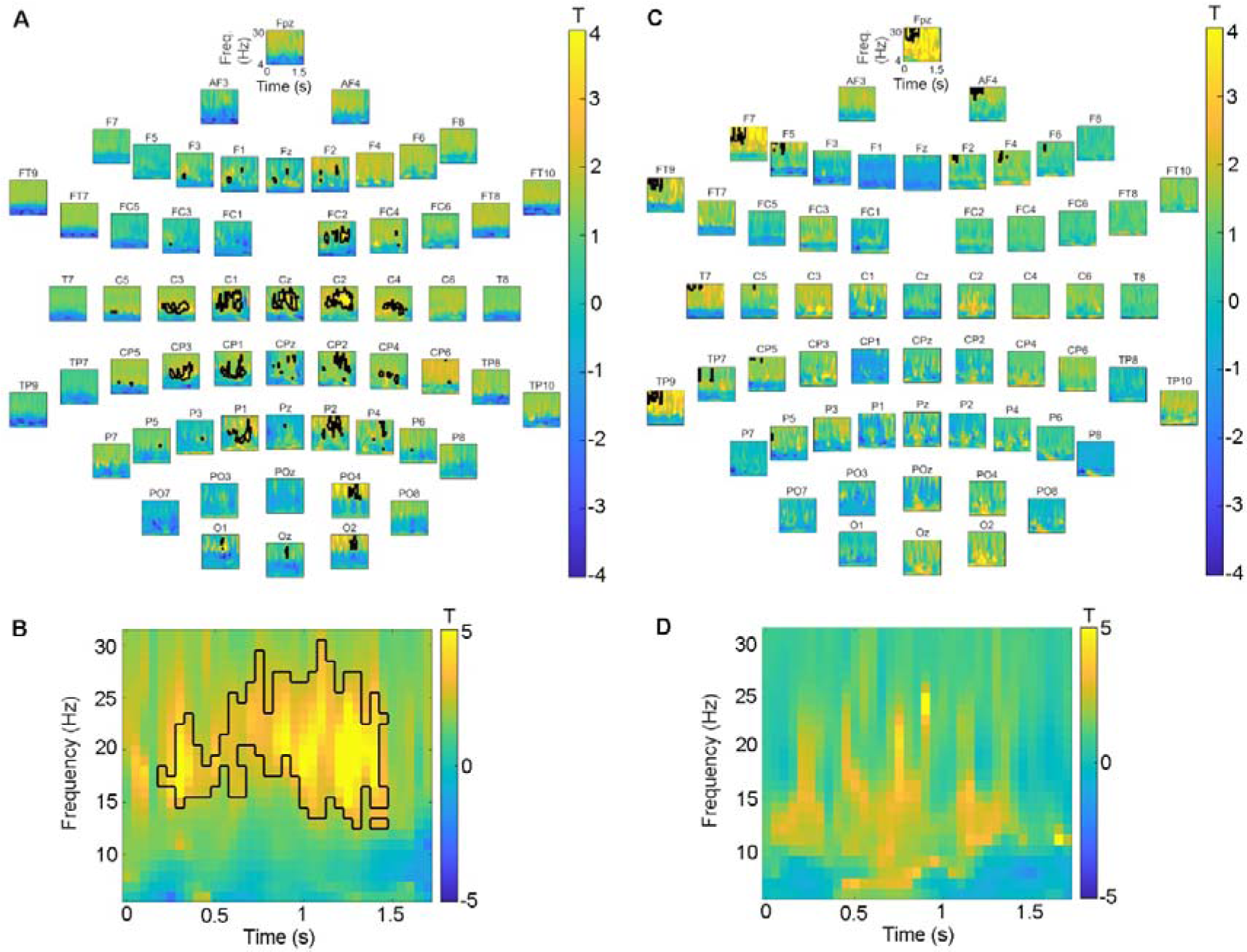
Spectral power pre-treatment minus power post-treatment in each group. Spectral power reduction was greatest from pre- to post-treatment over contralateral (ipsilesional) primary motor cortex in the BCI-FES group. Note that the positive T-values indicate a greater desynchronization post-than pre-treatment. Black contour = cluster of adjacent time–frequency points at which the post- vs-pre-treatment power differed according to paired T-tests at threshold p = 0.05. **A**. BCI-FES group: at each electrode. **B**. BCI-FES group: largest cluster observed at electrode C2, over right primary motor cortex. **C**. Random-FES group: at each electrode. **D**. Random-FES group: at electrode C2, over right primary motor cortex. Cluster-based permutation testing showed a significant difference between spectral power pre- and post-treatment in the BCI-FES group (p = 0.036).

**Figure 6.**
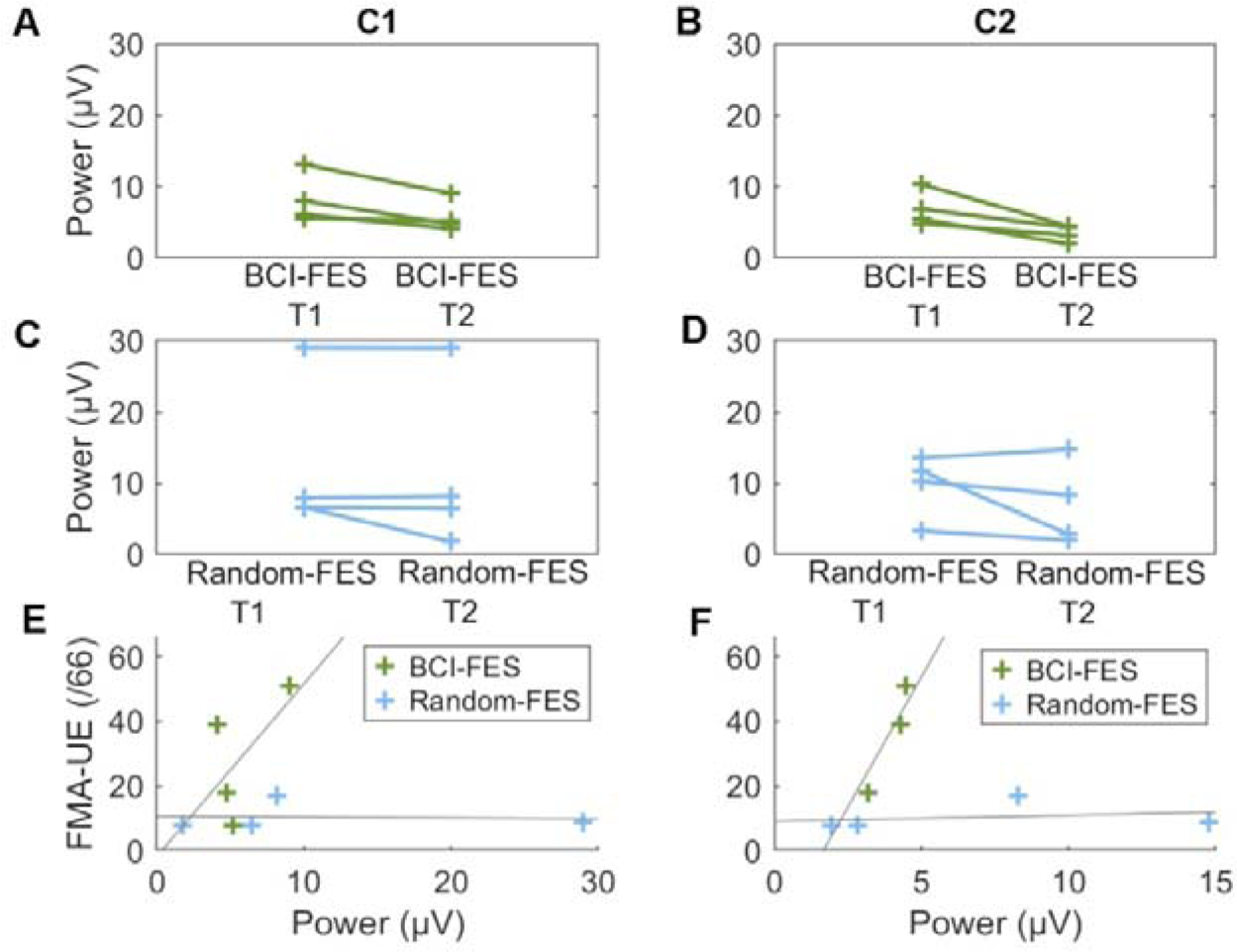
Individual patient beta (15-23 Hz) spectral power at 1.2 to 1.4 s post-movement cue before and after treatment. **A**. Over motor cortex ipsilateral to affected hand movement (C1) in the BCI-FES group. **B**. Over motor cortex contralateral to affected hand movement (C2) in the BCI-FES group. **C**. Over motor cortex ipsilateral to affected hand movement (C1) in the Random-FES group. **D**. Over motor cortex contralateral to affected hand movement (C2) in the Random-FES group. **E**. Correlation between beta spectral power ipsilateral to affected hand movement (C1) and FMA-UE after treatment. **F**. Correlation between beta spectral power contralateral to affected hand movement (C2) and FMA-UE after treatment (r(2) = 0.96, p = 0.044). No other correlation was significant.

LRTC, quantified using the Hurst parameter, was lower after than before the treatment program in the BCI group in the beta frequency range according to pairwise T-tests (Fig. 6A, B). Averaging over the beta frequency range at which power changed over time in the BCI-FES group (15-23 Hz) and over time, a reduction in LTRC was seen in the BCI-FES group only (paired T-tests, BCI-FES: T = −3.38, p = 0.043; Random-FES: T = 0.19, p = 0.86). While LRTC was higher after than before the program in the Random-FES group in the alpha frequency range (8-12 Hz), averaging over frequency and time, the difference was not significant (paired T-tests, BCI-FES: T = −0.85, p = 0.46; Random-FES: T = 0.52, p = 0.64). The reduction in beta-LRTC was consistently observed at an individual patient level in the BCI-FES group only, and the increase in alpha-LRTC was consistently seen in the Random-FES group only (Fig. 7C, D).

**Figure 7.**
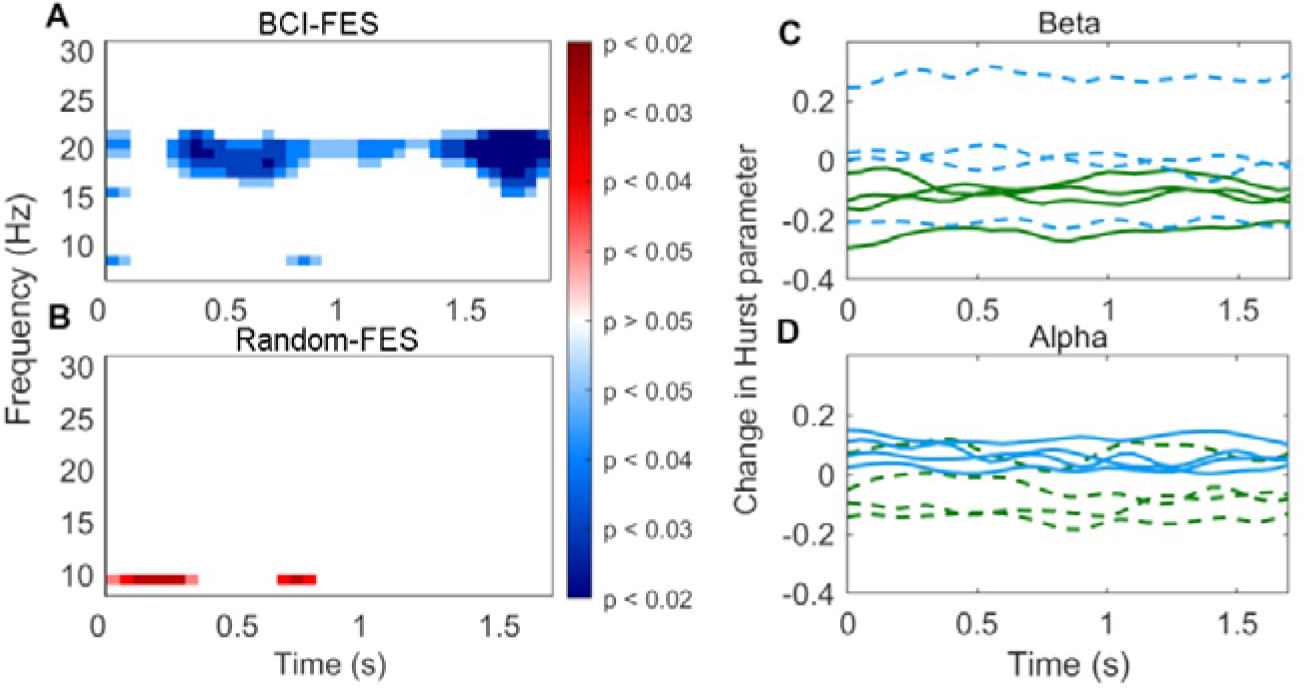
Changes in long-range temporal correlation (LRTC), quantified using the Hurst parameter, in high density EEG data recorded after compared with before the therapy program. Averaging over the beta frequency range at which power decreased post-therapy in the BCI-FES group (15-23 Hz) and over time, LRTC decreased only in the BCI-FES group (paired T-tests, BCI-FES: T = −3.38, p = 0.043. Random-FES: T = 0.19, p = 0.86). **A**, **B**. Significance of the pre- to post-treatment LRTC difference over frequency and time based on pairwise T-tests. **A**. BCI-FES group. **B**. Random-FES group. **C**, **D**. Changes in Hurst parameter in individual patients. Green: BCI-FES group; Blue: Random-FES group; Solid lines: significant difference on T-test in this group and frequency; Dashed lines: difference not significant **C**. At beta (18 Hz). **D**. At alpha (9 Hz).

We examined EEG–EMG coherence in the time–frequency window in which spectral power changed from pre- to post-treatment in the BCI-FES group (0.5-1.5 s; 15-23 Hz), at the electrode location over the contralateral primary motor cortex at which the power difference was greatest (C2). The EEG– EMG coherence was greater after than before treatment in the BCI-FES group (paired T-test: T = - 3.45, p = 0.041) but not in the Random-FES group (T = −0.073, p = 0.95) (Fig. 8).

**Figure 8.**
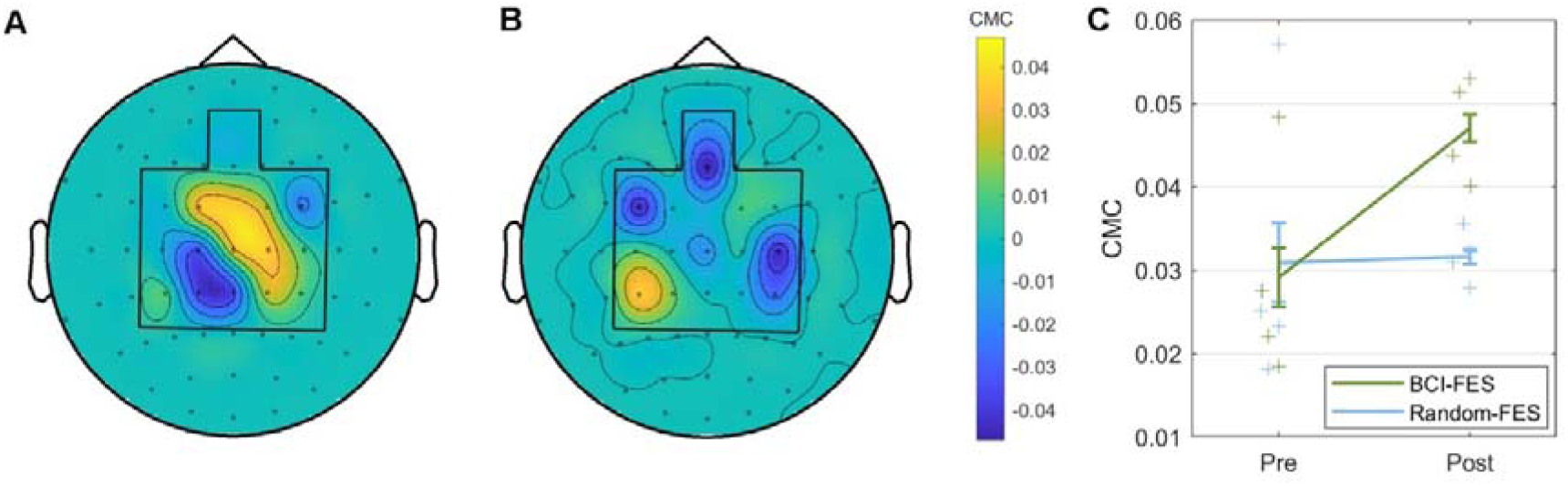
EEG–EMG coherence in the beta frequency range (0.5-1.5 s; 15-23 Hz): difference between pre- and post-therapy. **A**. BCI-FES group (paired T-test: T = −3.45, p = 0.041). **B**. Random-FES group (paired T-test: T = −0.073, p = 0.95). **C**. On an individual patient level.

## Discussion

Greater motor recovery, reflected by improved FMA-UE scores, was seen in the group receiving BCI– FES, with stimulation temporally locked to oscillatory spectral power changes in the sensorimotor rhythm, compared to the Random-FES group, who received FES at times unrelated to oscillatory correlates of movement attempts. Recovery was greater if the BCI–FES therapy was started in the acute phase post-stroke. Neural correlates of improved functional connectivity between contralateral (ipsilesional) motor cortex in the BCI-FES group included greater increases in TMS-induced MEP amplitudes and in corticomuscular coherence in the beta frequency range pre- to post-treatment than in the Random-FES group. Moreover, movement-associated beta spectral power reduction was more pronounced post-treatment in the BCI-than the Random-FES group, commensurate with a reduction in compensatory activity. Finally, long-range temporal correlation within beta oscillations was also reduced post-treatment in the BCI-FES group, suggesting that a subcritical state could be advantageous to motor recovery. Our findings are consistent with the proposal that FES delivery in a tight temporal window coupled with movement attempts using a BCI could improve post-stroke motor recovery, particularly if started early. Multiple neural correlates of motor recovery were modulated by the treatment program in the BCI group, supporting the notion that timing FES delivery according to sensorimotor electrophysiological correlates of movement attempts could have a specific impact on recovery processes. A strength of the study is the homogeneity of the patient group in terms of stroke location and laterality.

Few studies have investigated the potential impact of using BCI-FES early post-stroke^13,39,40^. Of the eight patients receiving BCI-FES in a partial crossover design study, four commenced treatment in the subacute phase, from 2-6 months post-stroke, with three in the BCI-FES and one in a control group receiving no FES^40^. Handedness, hemisphere affected, and lesion location varied. All three BCI patients showed improved motor function after treatment, while the control patient, whose impairment was also the most severe, did not. In another study with a partial crossover design, in which five of the 21 patients (mainly with stroke affecting the non-dominant hemisphere, including subcortical and cortical stroke) commenced treatment in the subacute stage, a clinically relevant improvement was seen in three of these patients^13^. A further study, involving seven right-handed patients with mainly subcortical stroke receiving BCI-FES in the acute/subacute phase, also showed greater motor recovery and enhanced sensorimotor rhythm desynchronization on the affected side after BCI-FES, which was not observed in the control group receiving FES unrelated to EEG features^39^. The improvements following BCI-FES in patients early post-stroke in these studies are consistent with our findings. Moreover, our preliminary analyses indicated that the increases in FMA and in beta desynchronization, as well as the reduction in beta LRTC, were observable in all four BCI-FES group patients individually^41^.

Similar to other studies applying BCI-based stroke rehabilitation, especially when starting early post-stroke^7^, our final sample size was small. This is a common problem in post-stroke rehabilitation studies^42^. The challenge of assigning comparable patients to large cohorts, due to the many patients who do not complete the treatment program, particularly when started early post-stroke, and the diversity of stroke location, highlights the importance of meta-analyses combining the findings from different studies. Meta-analyses include studies that fulfill the study evaluation criteria, including valuable data from studies with small sample sizes^7,43^, ranging from four to ten^44–51^, in which stroke type, laterality, location, and/or time after stroke varied within the sample. Providing data on an individual level from twelve patients in the current study will enable their integration in meta-analyses, with the additional advantages that the study group is homogenous with respect to important stroke features, with all patients having a subcortical stroke affecting the non-dominant hemisphere. Moreover, allocation of the patients to the intervention and control groups included counterbalancing in a randomized, double-blind study design according to age, sex, stroke type (ischaemic, haemorrhagic), and pre-treatment FMA.

Comparing alpha and beta oscillatory power pre- and post-treatment showed an increase in ipsilesional beta desynchronization in the BCI-FES group. On the other hand, alpha power provided more ipsilesional classification features by the end than at the start of the treatment program. Enhanced ipsilateral beta and also alpha desynchronization on motor imagery have been reported following BCI-based neurofeedback training in subacute stroke patients^52^. Modulations of alpha and beta power have been postulated to enable selection of task-relevant neural assemblies, with separate roles proposed for alpha and beta oscillations during goal-directed actions^53^. A decrease in contralateral sensorimotor beta power in healthy participants on increasing action selection difficulty was suggested to reflect disinhibition of cortical regions engaged in determining movement parameters, while increased ipsilateral alpha power was proposed to facilitate disengagement of task-unrelated neuronal populations^53^. Reinforcing alpha modulation associated with movement attempts, through providing visual and proprioceptive feedback generated by BCI-FES-induced movement using alpha power as a classifier feature, could have facilitated synaptic strengthening or maintenance of neuronal networks oscillating in the alpha frequency range involved in movement generation. Reducing the selection of beta power features for classification could have reduced the integrity of networks oscillating in the beta range. We note that greater pre-treatment ipsilesional alpha desynchronization has been associated with better outcome in chronic stroke patients, with increased desynchronization over a BCI-training program correlating with greater motor recovery^21^. A pre- to post-treatment change in movement-related sensorimotor oscillations in the BCI-FES group here is consistent with a modulatory effect of BCI-FES on the sensorimotor rhythm.

We observed a reduction in LRTC in beta oscillations in the BCI-FES group post-compared with pre-treatment. LRTC has been proposed to reflect neuronal systems close to a critical state, allowing fast reorganization of functional neural networks in response to changing demands. Better performance in an attentional task has been found to be associated with lower beta LRTC than at rest, and it was postulated that performance in tasks requiring sustained attention benefits from a sub-critical state^54^. LRTC in alpha band oscillations is also reduced following perturbation by a stimulus and on movement^34,55^. LRTC was not examined in the beta band in these studies, however. Our finding of reduced LRTC post-treatment in the BCI-FES group, who had shown better motor recovery than the Random-FES group, suggests moving to a sub-critical state is associated with improved motor function and could be induced by BCI-FES trained using the sensorimotor rhythm. Our findings are based on applying an ANOVA to the LRTC. We note that while permutation testing revealed only a trend towards a beta LRTC reduction in the BCI-FES group, examining the LRTC differences on an individual level indicates that each patient in the BCI-FES group showed a decrease in beta LTRC on an individual level, compared with only one patient in the Random-FES group.

Pre- with post-treatment comparison of electrophysiological markers differed on the contralateral (ipsilesional) side. An fMRI meta-analysis found that while contralesional motor cortical involvement is common, an eventual predominance of ipsilesional activity is associated with better motor outcome^33^. While lateralization of sensorimotor activity during post-stroke recovery to the contralesional hemisphere has been associated with better motor outcomes in a cohort including subcortical and cortical stroke patients^56^, better recovery has been reported with ipsilesional lateralization following subcortical stroke ^11,33,57^.

The main limitations of this study are associated with the early timing of the intervention and its impact on patient numbers. The most significant recovery post-stroke is seen in the first few weeks^3^, suggesting that intervention at this time may offer a window period with heightened neural plasticity, potentially enhancing facilitation of motor recovery. However, multiple factors contribute to the limited patient numbers included in BCI studies in early post-stroke patients^13,39,40^. Extensive investigations and treatments are frequently required on hospital admission, presenting a challenge to study recruitment. Moreover, co-existing medical conditions, often associated with the stroke, in this patient group can impede treatment program completion. Finally, spontaneous post-stroke recovery is most common in the acute phase, in the first days to weeks post-stroke^13^. These limitations are common across centers, underlining the need for multi-center studies and meta-analyses to address the efficacy of rehabilitation approaches in this group. Recruiting patients very early after stroke, while enabling our hypothesis to be addressed, is an important weakness in the study design, as these patients are more heterogenous in the extent of their spontaneous recovery than patients in the chronic phase post-stroke, making matched group allocation difficult without large patient numbers. The findings in the current study, given the low final sample size, should therefore be interpreted with caution, and be considered as providing a direction for further investigations rather than enabling firm conclusions regarding this rehabilitation approach.

The individual improvements in electrophysiological as well as clinical markers suggest that BCI-FES has the potential to be a promising approach to post-stroke rehabilitation. It is notable, however, that while the most marked motor recovery occurs in the acute phase, within the first 30 days post-stroke^58^, only a small improvement in FMA-UE score was observed in the patients starting the program in the acute phase in the Random-FES group. Greater improvement was seen in the Random-FES group in the subgroup starting in the subacute phase. An MRC score under 3 is predictive of poorer motor recovery^59,60^. By including this as an inclusion criterion, the expected spontaneous recovery was less. While starting rehabilitation early post-stroke is associated with higher recovery rates, it is also associated with higher dropout rates^61^. Two patients discontinued participation in the study during the initial evaluation phase due to complete recovery, which might explain the low spontaneous recovery time in the group starting in the acute phase. We used the Frane algorithm^62^ to weight the pseudorandom group allocation to balance potential confounding factors across the groups, including the initial FMA-UE score, but the high expected recovery in the acute phase underlines the importance of future work with larger patient groups. Particularly our findings in the acute subgroup should be viewed with caution, as the low spontaneous recovery rate in the Random-FES group could reflect an unintended bias in the group allocation, despite our use of the Frane algorithm, given the small sample size. Sample sizes are a major limitation in studies evaluating post-stroke rehabilitation approaches, and combining data from multiple centers will be a crucial step in evaluating their potential.

The non-dominant hemisphere was affected in the majority of patients able to participate, due to aphasia being an exclusion criterion. The laterality of brain activity associated with movement depends on whether the dominant or non-dominant side is affected and the handedness of the patient. Group level statistical analyses comparing pre- and post-treatment activity required these factors to be uniform across patients. A tendency to use the non-dominant hand less may impede use-related spontaneous recovery, which could play a role in the benefits seen following BCI-FES in this patient group. Further studies directly comparing groups in whom the non-dominant and dominant hemispheres affected are needed, but again, the group sizes required will necessitate large-scale multi-center patient recruitment to reach the necessary patient numbers in each group.

Measuring the amplitude of an MEP induced by TMS is a frequently applied method for motor recovery evaluation^60^. However, the amplitude depends on multiple factors. Transmission of a TMS pulse depends on intact cortical and also spinal synapses, and a single TMS pulse triggers a high frequency activity cascade in multiple pathways in the cortical region to which it is applied, so that MEP amplitude can only reflect corticospinal excitability in a general sense^63^. While we show that a clinically recognized post-treatment evaluation measure improved after treatment, these complex relationships, as well as the low participant number, should be taken into account when interpreting our findings. Future work involving TMS protocols with varying stimulation parameters and combining TMS with EEG has the potential to deliver more specific information about corticomuscular activity associated with post-stroke motor recovery and the impact of BCI-FES^63,64^.

We analyzed changes in MEP amplitude over the rehabilitation period, in response to TMS application at location C4 in all patients for whom these data were available from routine clinical assessment. We observed an interaction between group and time. However, the permutation tests showed a significantly higher MEP amplitude after than before treatment in both groups, although the degree of increase was significantly greater in the BCI-FES group. On an individual level, two of three BCI-FES group patients showed an increase, compared with one patient, with a smaller increase, in the Random-FES group. While sample size is an important consideration, given the small number of patients for whom MEP data were available, the approach taken to evaluating TMS should also be considered. C4 is considered to correspond with the hand area of the primary motor cortex^65,66^, and stimulation of C3/C4, with MEP measurement, is a standard evaluation approach^67–69^. While the precise, individual location of the motor hotspot corresponding to a particular muscle varies^66,70^, particularly after stroke^71^, measuring according to a fixed anatomical location allows a direct within-subject assessment of a change in MEP amplitude between time points. It is possible, however, that increased functional connectivity in patients in the Random-FES group was missed due to a change in location in the motor area corresponding with the electrode position over the extensor digitorum communis muscle. While the same operator performed all measurements, variability in coil placement could also affect MEP detection. Hotspot location determination can be optimized through combining TMS with neuroimaging and electrophysiology measurements^72^. Current work developing a stereotactic approach to motor mapping, based on individual neuroimaging and electrical field modelling, will provide more precise evaluation of motor recovery, including evaluation of network effects^73^. Although care was taken to match the groups according to lesion, specific lesion location also impacts wider network connectivity, and disconnection patterns may vary considerably between individuals.^74^ Future studies including muscle-specific TMS evaluation, also in combination with the electrode locations at which oscillatory activity provides the best movement classification for BCI-FES timing, would enable a more precise evaluation of the effects of BCI-FES on motor recovery.

Our findings support the proposal that using a BCI to trigger FES temporally coupled with movement attempts detected in motor cortical oscillations enhances post-stroke motor recovery, especially starting early after stroke. The electrophysiological findings suggest BCI-driven FES supports re-establishment of movement-associated processing on the ipsilesional side and a transition towards a subcritical state as contributing to the mechanism of Hebbian facilitation. Given the small sample size, however, further studies are required with larger numbers of patients to allow firm conclusions to be drawn.

## Methods

### Patients

The patients were a subgroup of the Magdeburg patient cohort in an international, multi-center double-blind, randomized controlled study, which comprised two registered trials with the same study protocol but differing target patient populations. The first trial targeted patients in the acute phase post-stroke (German Clinical Trials Register: DRKS00007832) and the second included patients in the subacute phase (DRKS00011522). Patients were recruited following acute hospital admission post-stroke or on transfer to the rehabilitation center, from the University Hospital Magdeburg stroke ward and the Neurorehabilitation Centre, MEDIAN, Magdeburg, Germany, respectively. The study protocol was approved by the Local Ethics Committee of the University Hospital, Magdeburg, Germany and performed in accordance with the principles of the Declaration of Helsinki. All patients discussed study participation and the possibility of withdrawing from the study at any time, without a need to provide a reason, with CMSR, and subsequently provided informed, written consent to participation.

#### Inclusion criteria

The primary inclusion criterion was upper limb paresis following stroke affecting wrist extension, with a Medical Research Council (MRC) Power Test score < 3, persisting >24 hours, and still present on recruitment. An MRC score of < 3 was chosen to focus on patients with lower chances of spontaneous recovery^59,60^. The acute group was recruited less than 1 month and the subacute group 1-6 months after stroke onset. Patients were required to be a minimum of 18 years of age, with no upper age limit. Diagnosis was confirmed using magnetic resonance imaging (MRI) or computerized tomography (CT), and patients with thrombotic or haemorrhagic stroke were included.

#### Exclusion criteria

The ability to understand the therapy instructions was a prerequisite, both to fulfill the requirement of provision of informed, written consent, and to enable active participation. Exclusion criteria were therefore a score < 25 on the Montreal Cognitive Assessment^75^ or severe aphasia, precluding active discussion of the instructions. Further exclusion criteria were severe hemi-neglect, depression (Hospital Anxiety and Depression Scale: HADS-total >15/21)^76^, fatigue (Fatigue Severity Scale > 36/63, i.e., > 4/7 on 9 items)^77^, pain in the neck/shoulder/arm (Pain Scale > 5/10)^78^, or a history of epilepsy. Other exclusion criteria were medical instability (orthostatic hypotension, sepsis, end-stage renal failure, severe visual impairment, fixed joint contractures, a skin condition that could be worsened through electrode placement), and taking certain regular medication (L-dopa, amantadine).

### BCI-FES

#### Group allocation

On recruitment, patients were pseudorandomly allocated to the BCI-FES or Random-FES group. The groups were counterbalanced according to the following factors: *Age*, *Sex*, *Lesion Side*, *Lesion Site* (subcortical, cortical), *Lesion Type* (ischaemic, haemorrhagic), and *Pre-treatment FMA*, to control for potential confounding factors. Patients were added sequentially to the database containing these factors and also the factor *Group Allocation*. The first four patients were allocated to the BCI-FES group, so that FES delivery parameters would be available for generating comparable parameters for the Random-FES group. Frane’s algorithm^62^ was then applied to the database to determine group allocation. An index of imbalance of each factor among patients so far recruited was calculated, based on each possible group allocation for the next patient. The index was a p-value from testing the hypothesis that the factor did not differ between groups. The Chi-square-goodness-of-fit test was used for *Group Allocation*, the Wilcoxon rank sum test for *Age* and *Pre-treatment FMA*, and the chi-square test for the remaining factors. For each possible group allocation, the largest imbalance was selected and converted to a probability of *Group Allocation* to each group by normalization. With each patient allocation, the most unbalanced factor at that time point was thus considered. The patients, therapists, and evaluating clinicians were blinded to group allocation.

#### EEG for the classifier

Sixteen EEG electrodes were placed bilaterally over motor cortical regions using a customized electrode cap, with electrode positions based on the 10-20 international system as follows: Fz, FC3, FC1, FCz, FC2, FC4, C3, C1, Cz, C2, C4, CP3, CP1, CPz, CP2, and CP4. The reference electrode location was the right mastoid, and the ground electrode was at AFz. Selective electrode coverage was used, as our aim was to base FES timing on motor cortical activity, and the reduced electrode number enabled rapid application, which was important for daily electrode application, to minimize therapist time and maximize compliance. EEG signals were recorded at a sampling rate of 512 Hz using a g.USBamp V2.14.07 amplifier (g.tec, Austria).

#### Therapy sessions

Patients received a maximum of five sessions per week, each occurring on different days. The total number of sessions depended on the length of the patients’ stay at the rehabilitation center. All patients received a minimum of three weeks, and a two-week extension was granted in certain cases by the individual state or private health insurance company. A mean of 18.8 [SD 5.7] treatment sessions were performed. Due to the variation, analyses of clinical outcomes were corrected for the number of sessions.

An initial training session was carried out to record EEG data during attempted movement and at rest, which were used to train the classifier. Patients were seated comfortably in front of a computer screen, with a table in front of them on which to rest their forearms, palms down, with flexed elbows. When a green up-arrow was presented, patients were instructed to attempt to extend the wrist of the paretic limb. To provide analogous visual stimulation for both trial types, a red down-arrow was presented when patients were to remain at rest. An upwardly moving bar was presented as visual feedback during movement attempts, and a downwardly moving bar was present during rest. The cue to begin each trial was presented at 0 s. Four to six five-minute blocks were performed.

Feature selection and classification were performed as in the previous chronic stroke study^12^. Following Laplacian-based spatial filtering, the Welch periodogram was applied to calculate the power spectral density at each electrode in 2 Hz bands from 8-30 Hz in 1 s sliding windows, shifting at 62.5 ms intervals (i.e., 16 times per second). Canonical variates analysis was used to identify up to 10 features for initial classifier training^79^. The trials were labelled as movement attempt or rest to provide input to train the Gaussian classifier using gradient-descent supervised learning. During the therapy, the probability was determined that a particular power spectral density value belonged to the movement attempt or rest trial class. When the classification threshold was not exceeded, a leaky integrator was used to smooth the ongoing output of the classifier. FES was triggered at the time point at which the probabilities integrated over time reached a threshold. If neither class was determined over a maximum 7 s trial, the trial was terminated, and the next one started. EEG data recorded during the therapy sessions were used to retrain the classifier each week, to account for changes over the course of the treatment.

Each subsequent therapy session comprised 3-7 blocks, according to fatigue levels, and lasted 10-25 minutes, including breaks. Fifteen movement attempts were made per block. For each therapy session, the EEG electrode cap was again applied, and two stimulating electrodes were placed over the extensor digitorum communis of the paretic forearm for inducing or assisting wrist extension by applying FES using a RehaStim stimulation device (Hasomed, Germany). EEG data were recorded continuously, with online classification 16 times per second. When a movement attempt was detected, FES was delivered. The Random-FES group had the same external set-up at the BCI-FES group, to enable blinding to group allocation, which is commonly referred to as “sham” treatment. To balance the stimulation frequency between the groups, a BCI-FES group patient was arbitrarily selected for each Random-FES group patient, and the corresponding frequency of stimulation was applied as a playback of that delivered to the BCI-FES group patient. This procedure ensured that the groups only differed in that the timing of FES in the Random-FES group was independent of the patient’s own cortical activity.

### Clinical evaluation

Clinical evaluations to compare the groups included direct physical assessment and impact on ability to perform daily tasks. The Edinburgh Handedness Inventory (EHI) was used to evaluate handedness.

The Fugl-Meyer Assessment upper extremity (FMA-UE) score (max. 66)^80^ was the primary outcome measure. A repeated measures ANOVA with the between-subject factor *Group* (BCI-FES, Random-FES), the within-subject factor *Time* (pre- and post-treatment), and the covariates *Age*, *Sex*, *Days Post-Stroke*, and *Days of Therapy* (i.e., number of treatment sessions) was used to compare the difference between FMA-UE score changes over the program between the groups. A repeated measures ANOVA was also applied including *Therapy start* (acute, subacute) as an additional between-subject factor.

We applied ANOVAs to clinical as well as electrophysiological markers, as they enable account to be taken of potentially important covariates as well as assessment of potential interactions. Using small sample sizes, T-tests and ANOVA are considered to have low statistical power, however, and it is challenging to prove the requirement of normal distribution. Simulation has provided support for the validity of T-tests with sample numbers as low as N = 2 to 5^81^, and as a generalization of the T-test, ANOVA is also applied to small sample sizes^82^. Moreover, no lower limit for sample size has been established for ANOVAs, but rather the key issue identified in considering sample samples sizes is whether they are representative of the studied population^83^. An important strength of the current study is the homogeneity of the patient group, supporting potential representativeness. However, given the small number of patients included in these analyses, we also provide individual data points in the figures to improve the interpretability of the results. Furthermore, we additionally applied two-sided permutation tests with 200 randomizations to make pairwise comparisons of mean values of each measure before and after the treatment program (see Supplementary information).

A range of secondary endpoints was determined, to enable a detailed exploration of any potential differences between the groups. They included the Medical Research Council Power Test, the Rivermead Test, the Barthel Index, the National Institute of Health Stroke Scale (motor: Arm), the European Stroke Scale, the Modified Ashworth Scale (spasticity), the Goal Attainment Assessment, and the Stroke Impact Scale.

### TMS

TMS was performed as a part of routine clinical monitoring from patients who fulfilled the inclusion and exclusion criteria relating to high magnetic field exposure. Before and after treatment, TMS was delivered to EEG location C4, over the primary motor cortex, while EMG was simultaneously recorded over the affected (left) extensor digitorum communis. TMS was commenced at 70% of capacity and increased repeatedly by 10%, until the maximum MEP amplitude was observed. The change in MEP from before to after treatment was compared between groups using a repeated measures ANOVA, with the between-subject factor *Group* (BCI-FES, Random-FES) and the within-subject factor *Time* (before, after), correcting for the covariates, *Age at stroke onset*and *Sex*.

### High-density EEG

#### Data recording

High-density EEG data were recorded using a BrainAmpDC amplifier (Brain Products GmbH, Germany) from 64 channels (sampling rate: 500 Hz), simultaneously with EMG data from electrodes placed over extensor digitorum communis of the affected limb during movement attempts, in twelve runs pre- and post-treatment. Each run comprised 10 movement and 5 rest trials in a pseudorandom order. Trials were presented using Presentation software (Version 18.2, Neurobehavioral Systems, Berkeley, CA, USA), analogously to movement cue presentation during the treatment program. The data were analyzed using custom Matlab scripts, EEGlab^84^, and FieldTrip^85^. Consistent with the clinical analyses, EEG data were analyzed from the patients with a non-dominant hemisphere, subcortical stroke. To enable electrode level comparison, we focused on patients who were purely right-handed (N = 8; BCI-FES: n = 4, Random-FES: n = 4).

#### Pre-processing

A notch (49–51 Hz) and a bandpass (1-200 Hz) filter were applied. The channels were then visually inspected and marked for ocular, EMG, and other artifacts. If >10 % of the data in a given channel were marked, it was replaced by spline-interpolated data from neighboring channels. The data were then re-referenced to an average reference, then epoched according to movement cue presentation (at time = 0 s) with a window of −2 s to 2.998 s (2500 frames). Epochs containing artifacts, determined by visual inspection, were excluded from subsequent analysis by JK and RK, supervised by CMSR. Independent component analysis (ICA) was applied, and components containing eye-blink, eye movement, and muscle artifacts were identified by JK and RK and removed, followed by back-projection of the ICs to the electrode space. The EMG data were epoched with the EEG data but separately notch- and high-pass filtered (10 Hz cut-off), then rectified. The data were further epoched to the times relevant for the subsequent analyses.

#### Spectral power analysis

Time-frequency decomposition was carried out through convolution with 5-cycle Morlet wavelets from 4 to 31 Hz. Change in oscillatory spectral power from pre- to post-treatment was compared for each group. Paired T-tests were applied to each time-frequency point, with a threshold of p = 0.05, followed by cluster-based permutation tests with 500 randomizations. We then examined the change in individual patient beta spectral power pre- to post-treatment on an individual level over motor cortex ipsi- and contralateral to movement of the affected hand for each group, followed by calculation of Pearson’s correlation coefficient between post-treatment contralateral beta spectral power and FMA-UE.

#### Corticomuscular coherence

Coherence was calculated between the EMG signal recorded over the extensor digitorum communis during movement attempts and each EEG channel in the time–frequency window (0.5 to 1.5 s, 15-23 Hz) at which the pre- to post-treatment spectral power reduction differed between the BCI- and Random-FES groups. The EMG and EEG data were Fourier-transformed, with multitaper spectral smoothing, and the cross spectra were calculated based on the phase difference between the EMG and each EEG signal. The change in EEG–EMG coherence from pre- to post-treatment was compared for each group over contralateral motor cortex, at electrode C2, where power modulation was greatest, using paired T-tests.

#### Long-range temporal correlation

LRTC was calculated using detrended fluctuation analysis (DFA). DFA was developed, because autocorrelation function analyses may yield spurious long-range correlations when the data are non-stationary. Evaluation of the decay in auto-correlation between remote parts of a non-stationary data sequence using DFA^86^ is therefore applicable in EEG data^34^. LRTC can be quantified in EEG data in either the time or the frequency domain, the former by fitting the power law to the autocorrelation, and the latter by estimating the slope of the 1/f power spectrum on a log–log scale and computing the scaling exponent. DFA provides a more practical and most common approach to quantifying the degree of temporal dependency in non-stationary signals, captured in the Hurst exponent (H), and has been shown to be consistently related to both of those approaches^87^. In EEG signals, the degree of self-similarity within the time series has previously been quantified based on power law scaling, by applying least squares linear regression to determine the slope of a log–log plot of detrended fluctuations against window size (time scale) to yield H^55,86,88^. LRTC is deemed present when H is between 0.5 and 1.

LRTC in alpha and beta oscillations partially overlaps topologically with the distribution of spectral power, and alpha and beta power and LRTC correlate weakly^35^. We therefore evaluated LRTC at the electrode location at which power differences from pre- to post-treatment differed most between the BCI- and Random-FES groups. The data were time–frequency decomposed using the wavelet transform with 5-cycle wavelets, amplitudes were extracted for alpha and beta frequencies (9-30 Hz), and H was calculated in 1 Hz steps. Long signal segments are needed to estimate H in narrowband signals^89^, so we concatenated the movement trials before applying DFA, following the approach of Wairagkar and colleagues^55^, as the DFA scaling exponent is not affected by stitching data together^88,90^. The minimum available number of trials for a given patient was 20, so 20 sequential trials were concatenated for each patient. The LRTC was then calculated over a 47.5 s sliding window in 50 ms steps, and the LRTC value was assigned to the first time point of each window. Paired T-tests were applied to compare LRTC before and after treatment for each group across time and the alpha and beta frequency ranges, as these frequencies were used as classifier features during the treatment program.

## Acknowledgements

The authors would like to thank Anne-Katrin Baum, Manuela Reichwald, and Angelika Klemme for support in data acquisition and Arne Leukert and Luise Ebeling-Jung for carrying out the therapy sessions. We also thank the patients for their participation.

## Author contributions statement

CMSR: wrote first draft of manuscript; JK, CR, SP, SV, TH, SD, RTK, HH, HJH, MS, JRM, CMSR: study concept and design; CMSR: study coordination; JK, RK, SV, TH, AS, JL, AH, MG, HJH, SL, MS, CMSR: patient recruitment; CMSR: patient consent; JK, RK, SV, TH, AH, SL, CMSR: clinical evaluation for inclusion; JK, RK, SV, TH, AH, SL: blinded pre- and post-therapy clinical evaluation; SV, TH, AH, MG, HJH, SL, MS: clinical management; AS, JL, AH, SL, MS, CMSR: therapy coordination; CR, SP, JRM, CMSR: group allocation, therapy preparation, and implementation; CR, CMSR: instructed therapists in treatment implementation; SP, JRM: wrote group allocation and classifier software; JK, RK, CMSR: electrophysiological recording; JK, RK, CMSR: electrophysiological and TMS analyses; JK, CR, SJN, ICT, RTK, CMSR: data interpretation. All authors approved the final manuscript.

## Data availability

The datasets analyzed during the current study are available from the corresponding author on reasonable request.

---

**Supplementary Figure 1.**
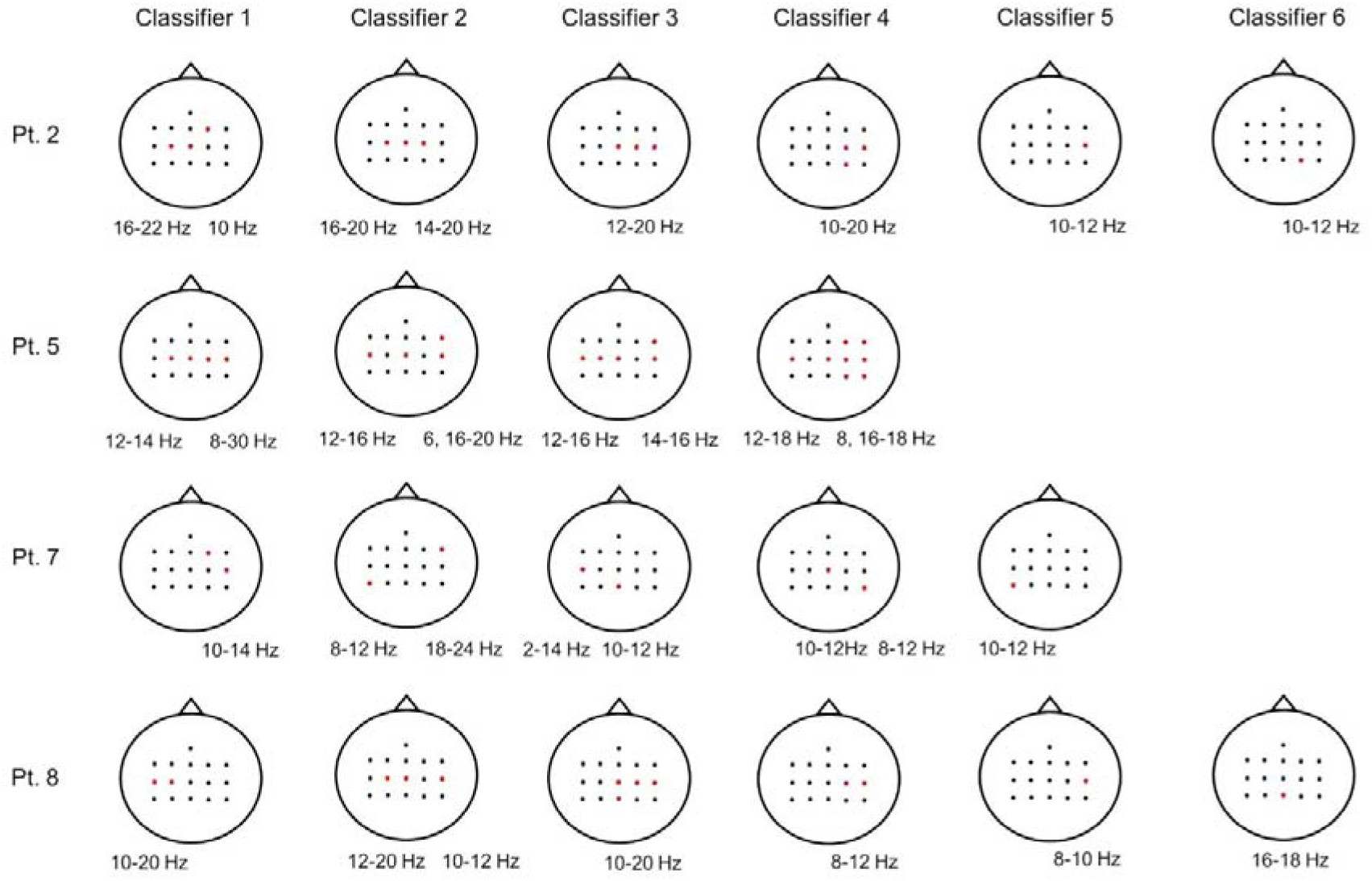
Classifier features. Features selected for classifiers for patients in the BCI-FES group over the course of the therapy program. Note that classifiers were not trained to determine the timing of functional electrical stimulation in the Random-FES group. Selected electrodes are highlighted in red, and the frequencies chosen are given below the relevant electrode.

